# Comprehensive analysis across *SMN2* excludes DNA methylation as an epigenetic biomarker for spinal muscular atrophy

**DOI:** 10.1101/2024.11.21.24317551

**Authors:** M.M. Zwartkruis, J.V. Kortooms, D. Gommers, M.G. Elferink, I. Signoria, J. van der Sel, P.J. Hop, R.A.J. Zwamborn, R. Geene, J.W. Green, H.W.M. van Deutekom, W. van Rheenen, J.H. Veldink, F. Asselman, R.I. Wadman, W.L. van der Pol, G.W. van Haaften, E.J.N. Groen

**Affiliations:** Department of Neurology and Neurosurgery, UMC Utrecht Brain Center, University Medical Center Utrecht, Utrecht, the Netherlands; Department of Genetics, University Medical Center Utrecht, Utrecht, the Netherlands; Department of Translational Neuroscience, UMC Utrecht Brain Center; Center for Molecular Medicine, University Medical Center Utrecht, Utrecht, the Netherlands; Utrecht Sequencing Facility, Center for Molecular Medicine, University Medical Center Utrecht, Utrecht, the Netherlands; Oncode Institute, Utrecht, the Netherlands

**Keywords:** spinal muscular atrophy, survival motor neuron gene, DNA methylation, long-read sequencing, bisulfite sequencing

## Abstract

Spinal muscular atrophy (SMA) is a severe neurodegenerative disease caused by defects in the survival motor neuron 1 (*SMN1*) gene. The wide variability in SMA severity is partially explained by an inverse correlation with copy number variation of the second human *SMN* gene (*SMN2*). Nevertheless, significant variability in severity and treatment response remains unexplained, prompting a search for accessible biomarkers that could explain and predict this variability. DNA methylation of *SMN2* has been proposed as one such biomarker, but comprehensive evidence and analyses are lacking. Here, we combined long-read nanopore sequencing with targeted bisulfite sequencing to enable high-resolution analysis of *SMN2*-specific methylation patterns. We observed tissue-specific variation in DNA methylation across the entire 30 kb *SMN2* gene in 29 patients analyzed by long-read nanopore sequencing, identifying variable methylation patterns in the promoter, introns, and 3’ UTR. Subsequent targeted analysis of these regions by bisulfite sequencing of blood-derived DNA in 365 SMA patients showed no association between *SMN2* methylation and disease severity or treatment response, excluding blood methylation patterns as predictive biomarkers. However, we discovered significant age-associated variation in *SMN2* methylation, particularly in intron 1 and the 3’ UTR, highlighting DNA methylation as a possible modifier of SMN expression during development and aging. Our approach provides a broadly applicable strategy for detailed but cost-effective and high-throughput characterization of DNA methylation in other genes and diseases, including complex genetic regions.

## Introduction

Spinal muscular atrophy (SMA) is a devastating neurodegenerative disease characterized by progressive muscle weakness and atrophy that may cause infantile death or severe childhood disability. The incidence is approximately 1 in 6,000-10,000 live births [1]. SMA is caused by homozygous deletion or mutation of the *survival motor neuron 1* (*SMN1*) gene, which results in insufficient production of the critical and ubiquitously expressed SMN protein [2]. While SMA patients lack a functional *SMN1* gene, they retain one or more copies of the highly similar *SMN2* gene. The *SMN2* gene can partially compensate for the loss of *SMN1*, but its ability to do so is limited by alternative splicing that excludes exon 7, producing a truncated and unstable form of the SMN protein [3–5]. The number of *SMN2* gene copies, ranging from one to six in patients with SMA, inversely correlates with disease severity, with higher copy numbers associated with milder phenotypes [6,7]. However, this relationship is not absolute, and significant variability in clinical presentation exists even among patients with the same *SMN2* copy number [6,8]. Additionally, the expression of SMN is strongly developmentally regulated through unknown mechanisms, with high pre- and neonatal expression followed by a reduction later in life [9,10]. Finally, treatment response to the three currently available gene-targeted therapies varies between patients with equal *SMN2* copy numbers [11,12]. These observations suggest that factors beyond *SMN2* copy number may influence SMN protein expression and SMA outcomes.

One potential source of this variability may lie in the epigenetic regulation of *SMN2*, as has often been hypothesized [7,11,13–16]. DNA methylation is a key epigenetic mechanism that can influence gene expression without altering the underlying DNA sequence [17]. Previous studies have identified differential methylation patterns in the *SMN2* promoter region between SMA patients with varying disease severities, suggesting that DNA methylation may influence *SMN2* expression [18,19]. Furthermore, the methyl-CpG-binding protein MECP2 has been shown to interact with methylated sites in the *SMN2* promoter, potentially regulating *SMN* gene activity [18,20]. Several studies investigated genome-wide differential DNA methylation in SMA patients, however with limited sample sizes [21,22]. While these initial findings are intriguing, the existing literature on *SMN2* methylation in SMA is limited, often focusing on a small number of CpG sites within the promoter region and in small patient cohorts. Our overall understanding of variation in DNA methylation in patients with SMA therefore remains limited. To fully elucidate the role of DNA methylation in the regulation of *SMN2*, a more comprehensive analysis across the entire gene is warranted. In this study, we therefore aimed to provide a detailed characterization of DNA methylation patterns across *SMN2* in a large cohort of patients with SMA, using a combination of long-read nanopore sequencing and targeted bisulfite sequencing. We identified lowly methylated CpG sites in the *SMN2* promoter, the transcription site of the long non-coding RNA *SMN-AS1*, several intronic regions, and the 3’ UTR of *SMN2*, with tissue-specific differences. No association between DNA methylation and disease severity or treatment response was found, ruling out DNA methylation as an epigenetic biomarker of SMA. DNA methylation was significantly associated with age at the *SMN2* 3’UTR and intron 1, suggesting these changes might be involved in the regulation of SMN expression during development and aging.

## Results

### Nanopore sequencing reveals extensive variation in DNA methylation across SMN2 and copy-specific differential methylation between SMN2 haplotypes

To comprehensively explore DNA methylation across *SMN2* in SMA patients, we determined CpG methylation status from nanopore sequencing data from 10 blood samples and 22 fibroblast samples from 29 patients with varying *SMN2* copy numbers and SMA types (**Table S1**) [23]. We aligned the sequencing reads to *SMN1* by masking *SMN2* and phased the reads into two to five haplotypes as described previously [23]. We analyzed DNA methylation per patient and per *SMN2* haplotype (**Fig. 1A**) and visualized the methylation percentage per CpG site in a heatmap (**Fig. 1B**). In addition to low methylation of the promoter region, low methylation was also observed in the transcription site of the lncRNA *SMN-AS1* [14], the 3’ UTR and various other, intronic regions of *SMN2*. Hierarchical clustering (**Fig. 1B**) and principal component analysis (PCA, **Fig. 1C**) showed that methylation data from fibroblast and blood clustered separately, as expected based on previous reports showing variation in DNA methylation between tissues [24]. Fifty-eight sites were differentially methylated (p_adj_<0.01) between blood and fibroblasts (**Fig. 1B** and **1D**). Because of this, subsequent analyses were performed separately for each tissue. When exploring a possible correlation between DNA methylation and disease severity by SMA type in this small group of samples, no differentially methylated sites were found (**Fig. 1E-F**). We and others previously identified multiple single nucleotide variants (SNVs) that can be used to study the genomic environment of individual *SMN* copies and serve as markers of gene conversion [23,25] and used these insights to next study DNA methylation per *SMN* haplotype (**Fig. S1A**). When comparing *SMN2* haplotypes with *SMN1* environment SNVs to those with *SMN2* environment SNVs (**Fig. 1G**), we found five differentially methylated CpG sites in blood and seven in fibroblasts (p_adj_<0.01), of which four overlapped (**Fig. 1H-I, Fig. S1**). However, most of these are at known SNV positions [23] (**Fig. S1B-C**), and these differences are therefore likely caused by a nucleotide variant in the CpG site, resulting in a non-CpG site (**Fig. 1J**). Non-CpG methylation is possible in humans, but its properties and functions are not well known [26,27]. Hence, it was not included in the methylation calling algorithms of this study (**Methods**). Interestingly, although based on a limited number of observations, one of the differentially methylated sites in blood (chr5:71381516, promoter region) was not a known SNV site, indicating a possible association between gene environment and methylation status. In summary, ONT sequencing allowed us to study DNA methylation across the entire *SMN2* gene and led us to identify several regions of interest (ROIs) for further analysis in a larger cohort.

**Figure 1:**
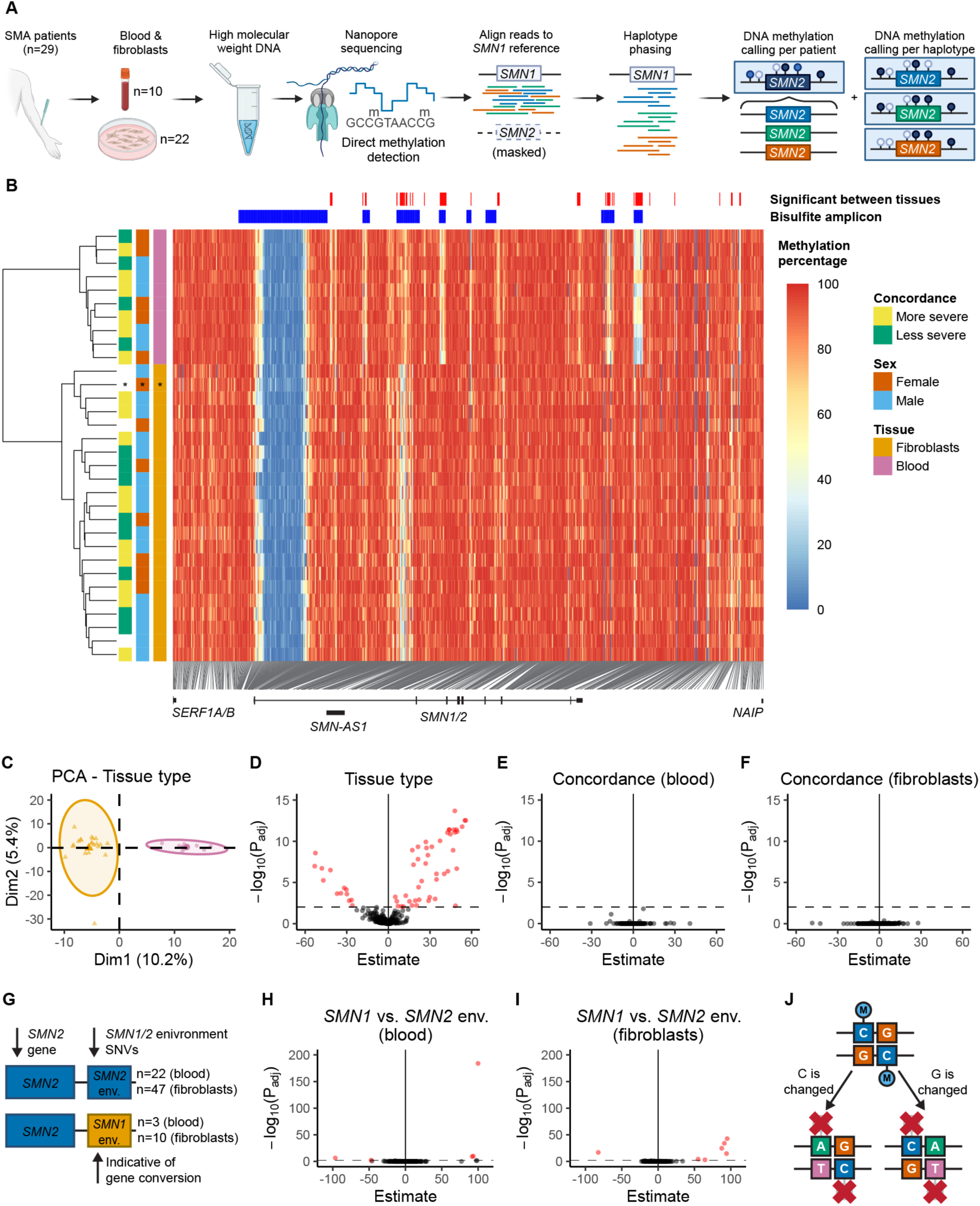
DNA methylation varies extensively throughout the *SMN2* gene and varies between tissues. (A) Overview of sequencing and bioinformatics approaches in this study. Nanopore sequencing with adaptive sampling was performed on high molecular weight DNA of SMA patients. Reads were mapped to *SMN1* and phased into haplotypes as described previously [23]. Methylation percentage per CpG site was determined per patient and per haplotype. Created with Biorender.com. (B) Heatmap of CpG DNA methylation in and around the *SMN2* gene (T2T-CHM13 chr5:71375000-71425000) in SMA patients with a homozygous *SMN1* deletion and one patient with one copy of *SMN1* with a pathogenic mutation (indicated with an asterisk). Each row represents one patient, each column represents one CpG site. Hierarchical clustering with the ward.D2 method was performed on the rows. Patient characteristics are shown left of the heatmap. (C) Principal component analysis (PCA) of the methylation data from (B). Blood samples (pink) cluster separately from fibroblast samples (orange). (D) Differential methylation analysis of DNA methylation between fibroblasts (n=22) and blood (n=10). 58 sites are differentially methylated between tissues (p_adj_<0.01). (E) Differential methylation analysis of DNA methylation between more severely (n=6) and less severely (n=4) affected patients for blood samples. No differentially methylated sites were found (p_adj_<0.01). (F) Differential methylation analysis of DNA methylation between more severely (n=10) and less severely (n=8) affected patients for fibroblast samples. No differentially methylated sites were found (p_adj_<0.01). (G) Schematic representation of haplotypes with *SMN1* environment SNVs and *SMN2* environment SNVs. (H-I) Volcano plot DNA methylation of haplotypes with *SMN1* environment SNVs versus haplotypes with *SMN2* environment SNVs, for blood (H) and fibroblasts (I). Five differentially methylated sites were found in blood and seven sites in fibroblasts. Sample sizes are as indicated in (G). (J) Schematic representation of a methylated CpG site, and likely consequences for methylation if either the C or G changes to another nucleotide: methylation is not present or not detected by the currently used algorithms.

### DNA methylation in the SMN2 gene can be determined by targeted bisulfite sequencing

To investigate whether variation in DNA methylation identified by ONT sequencing was linked to disease severity or treatment response, we performed targeted bisulfite sequencing on DNA isolated from blood from 365 SMA patients (**Fig. S2**, **Table 1, Table S2-3**). Per patient, we pooled 18 amplicons containing the ROIs identified by ONT sequencing (**Fig 2A**, **Table S4**), 15 of which were successfully sequenced by barcoded Illumina sequencing (>100x read depth in at least 90% of patients). We included only CpG sites that were mapped on the intended targets with at least 100x read depth and excluded potential SNV sites (**Fig. S3A**). Median read depth per CpG site was 3,014-37,870x and for every CpG site, 95-100% of the patients had a methylation call (**Fig. S4A-B**). For nine patients, both nanopore and bisulfite methylation data was available; methylation percentages showed high correlation between data types (Spearman’s R=0.77, *p<*2.2e-16, **Fig. S4C**). Heatmap visualization of methylation percentage per CpG site illustrated that most variation was present in the promoter region and 3’UTR. We performed hierarchical clustering of patients and found that clustering was mostly based on variation in intron 1 and the 3’ UTR of *SMN2* (**Fig. 2B**). We performed further dimensionality reduction of the dataset by filtering out CpG sites with little variation in methylation percentage (standard deviation <5%) and CpG sites with high correlation (Spearman R>0.9), resulting in 57 sites for further analysis and statistical testing (**Fig. S3A-B**).

**Figure 2:**
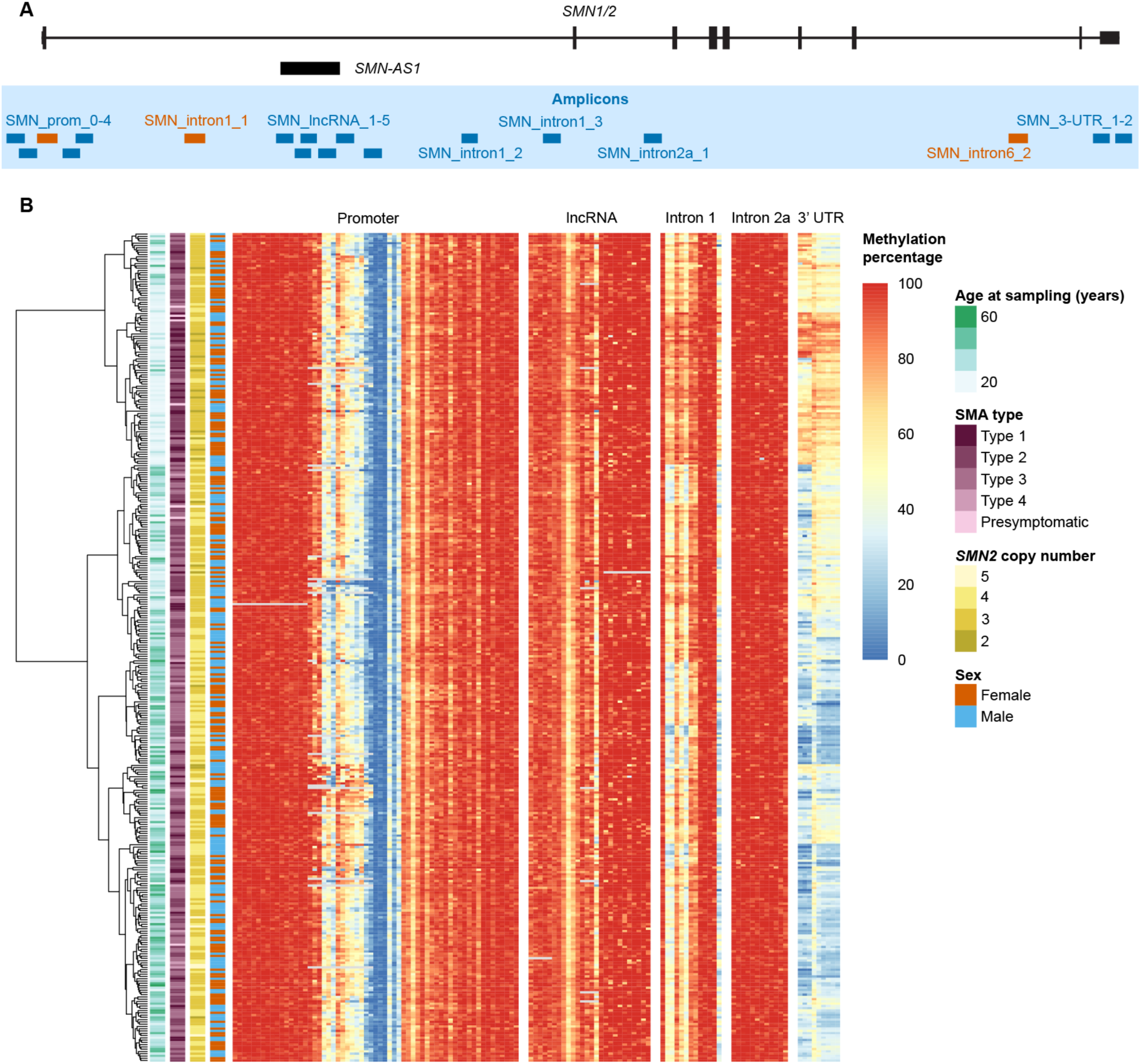
DNA methylation in the *SMN2* gene can be determined by targeted bisulfite sequencing. (A) Location of 18 amplicons, 15 of which were sequenced successfully (blue, a total of ∼6.6kb around *SMN2*). Orange indicates amplicons with insufficient coverage for further analyses. (B) Heatmap of DNA methylation percentage per CpG site determined with targeted bisulfite sequencing. Each row represents one patient, each column represents one CpG site. Hierarchical clustering with the ward.D2 method was performed on the rows. Patient characteristics are shown left of the heatmap. Clustering of patients is mostly based on DNA methylation in intron 1 and the 3’UTR.

**Table 1:** Baseline characteristics of SMA patients sequenced with Nanopore sequencing with homozygous *SMN1* deletion and no c.859G>C variant in *SMN2*.

|  | 2x*SMN2* | 3x*SMN2* | 4x*SMN2* | 5x*SMN2* | Overall |
| --- | --- | --- | --- | --- | --- |
| **Total** | 15 | 215 | 122 | 7 | 359 |
| **Sex** |  |  |  |  |  |
| Male | 10 (66.7%) | 94 (43.7%) | 71 (58.2%) | 6 (85.7%) | 181 (50.4%) |
| Female | 5 (33.3%) | 121 (56.3%) | 51 (41.8%) | 1 (14.3%) | 178 (49.6%) |
| **SMA type** |  |  |  |  |  |
| Type 1 | 15 (100%) | 30 (14.0%) | 1 (0.8%) | 0 (0%) | 46 (12.8%) |
| Type 2 | 0 (0%) | 138 (64.2%) | 14 (11.5%) | 0 (0%) | 152 (42.3%) |
| Type 3 | 0 (0%) | 43 (20.0%) | 96 (78.7%) | 5 (71.4%) | 144 (40.1%) |
| Type 4 | 0 (0%) | 0 (0%) | 9 (7.4%) | 1 (14.3%) | 10 (2.8%) |
| Presymptomatic | 0 (0%) | 4 (1.9%) | 2 (1.6%) | 1 (14.3%) | 7 (1.9%) |
| **Age at onset (years)** |  |  |  |  |  |
| Mean (SD) | 0.205 (0.153) | 1.08 (0.987) | 7.39 (9.45) | 11.5 (6.23) | 3.30 (6.35) |
| Median [Min, Max] | 0.208 [0, 0.500] | 0.917 [0, 9.50] | 2.50 [0.458, 43.0] | 12.5 [1.00, 19.0] | 1.08 [0, 43.0] |
| Missing | 1 (6.7%) | 11 (5.1%) | 13 (10.7%) | 1 (14.3%) | 26 (7.2%) |
| **Age at sampling (years)** |  |  |  |  |  |
| Mean (SD) | 3.57 (4.64) | 21.9 (16.9) | 35.5 (20.7) | 31.2 (14.4) | 25.9 (19.6) |
| Median [Min, Max] | 1.86 [0.282, 18.5] | 19.6 [0.0438, 64.2] | 37.3 [0.189, 79.7] | 25.0 [18.0, 52.3] | 23.1 [0.0438, 79.7] |

### DNA methylation significantly correlates with age at 22 CpG sites across SMN2

To explore whether DNA methylation was associated with baseline patient characteristics – such as sex and age – we performed PCA, excluding patients with uncommon genotypes: carriers of an *SMN1* copy with loss-of-function variants or the positive modifier c.859G>C in *SMN2*. Male and female groups completely overlapped (**Fig. 3A**), whereas age group clusters only partly overlapped (**Fig. 3B**). Differential methylation analysis did not yield differentially methylated sites between male and female patients (**Fig. 3C**). In contrast, DNA methylation was significantly associated with age at 15 sites including three collapsed sites (p_adj_<0.01, **Fig. 3D**), corresponding to 22 CpG sites in total. Interestingly, DNA methylation percentage increased with age at one site (promoter region) but decreased at all other significantly associated sites in intron 1, the 3’ UTR and the lncRNA *SMN-AS1* sequence (**Fig. 3E**). As this association implies that age may be a confounding variable, we corrected for it in our subsequent statistical analyses. The sites associated with age did not contain common nucleotide motifs (**Fig.S5**). Altered DNA methylation may affect binding of CCCTC-binding factor (CTCF), which may in turn affect 3D genome organization and alternative splicing [28–30]. However, CTCF binding site motifs (CTCF_HUMAN.H11MO.0.A on hocomoco11.autosome.org [31]) did not overlap with any of the age-related CpG sites in *SMN2* [32] (**Fig. S6**). To investigate whether age-associated changes in *SMN2* DNA methylation affect *SMN2* mRNA expression, we made use of previously generated expression data from whole blood [8]. We found that *SMN2-FL* RNA expression was significantly associated with age in patients with three copies of *SMN2*, but not in patients with four copies of *SMN2* (**Fig. S7A**), possibly because this group did not include patients younger than 6 years old and the sharpest decline of SMN expression happens perinatally [9]. We tested whether there was an association between DNA methylation and *SMN2-FL* expression in patients with three *SMN2* copies. An association can be observed between DNA methylation and *SMN2-FL* expression at most CpG sites associated with age, where methylation appeared to be higher in patients with higher *SMN2-FL* expression (**Fig. S7B**), although these associations were not statistically significant (**Fig. S7C**).

**Figure 3:**
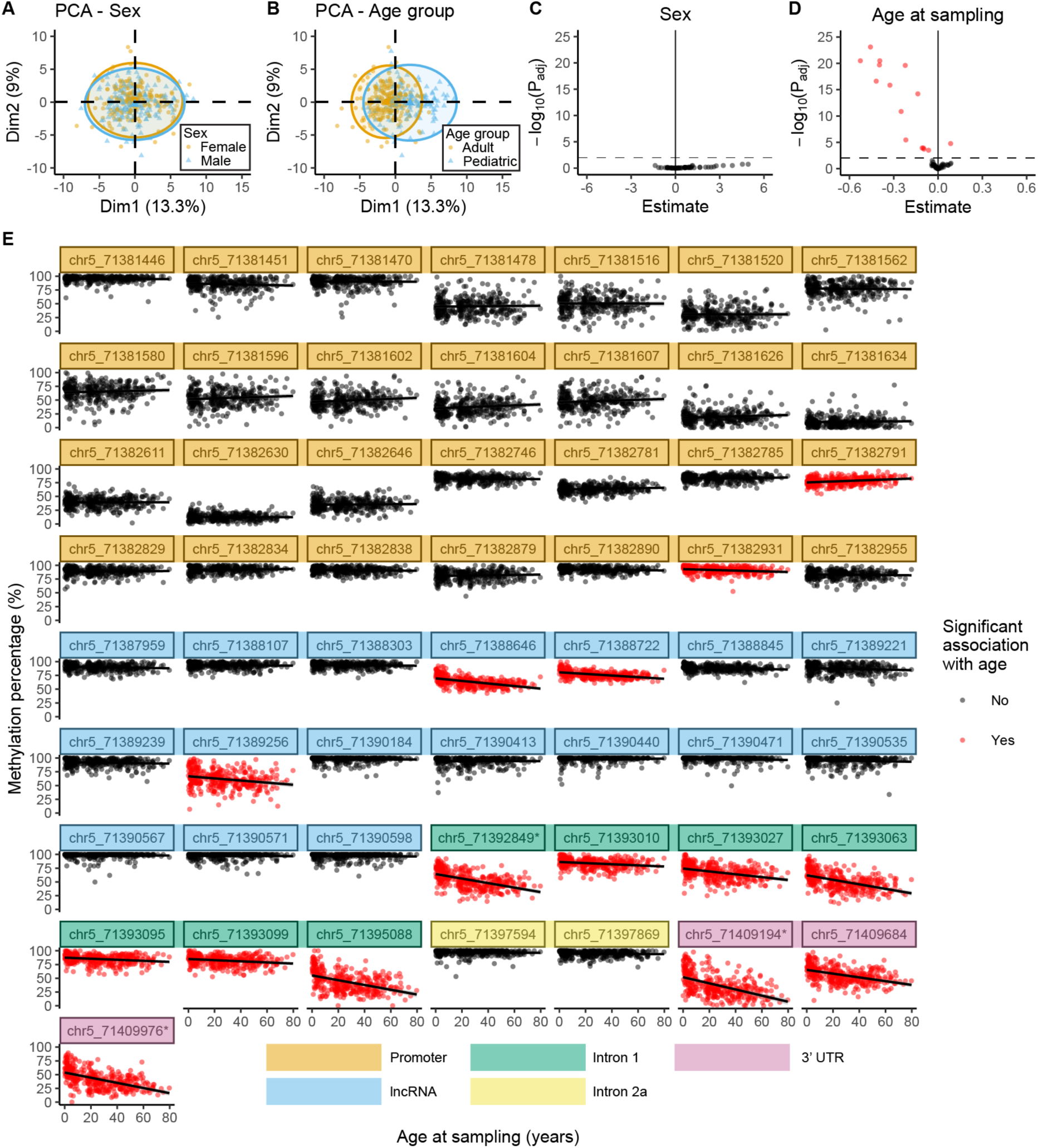
DNA methylation in *SMN2* is significantly associated with age at sampling. (A-B) Principal component analysis (PCA) of the methylation data used for statistical analysis, colored by sex (A) or age (B). Sex clusters overlap completely, whereas age group clusters overlap only partly. (C-D) Differential methylation analysis for sex (C) and age at sampling (D) shown with volcano plots (n=359). Fifteen sites including collapsed sites, corresponding to 22 CpG sites in total, are significantly associated with age (p_adj_<0.01). (E) Methylation percentage of all sites analyzed in the differential methylation analysis. Differentially methylated sites from (D) are colored red. DNA methylation significantly decreased with age in all tested sites in intron 1 and the 3’UTR, and several sites in the promoter and lncRNA and DNA methylation at one site in the promoter significantly increased with age. Sites that were collapsed for statistical analysis (see **Methods**) are indicated with an asterisk (*).

### No association between DNA methylation in SMN2 and disease severity or treatment response

To investigate whether DNA methylation was associated with disease severity or treatment response, we first analyzed patients with genetic variants that potentially modify disease severity separately. We found no association between DNA methylation and presence of *SMN1* or the c.859G>C variant in *SMN2* (**Fig. S8**). Patients with these genotypes were excluded from further analyses, since their phenotypes are likely affected by these genetic variants. To test if DNA methylation was associated with *SMN2* copy number, we first performed PCA that suggested there might be differences between copy number groups (**Fig. 4A**). However, this observation was likely confounded by age as mentioned above, since patients with lower *SMN2* copy number are often sampled at younger ages than patients with higher copy number. Indeed, when correcting for age, none of the tested sites were significantly associated with *SMN2* copy number (**Fig. 4C**). Similarly, no association was found between DNA methylation and copy number of the *NAIP* gene (**Fig. S9A**), which has also been suggested as a potential modifier of SMA, although inconclusively [13]. No significantly differentially methylated sites were found for disease severity by SMA type (**Fig. 4B** and **4D**) or age at onset (**Fig. S9B**). No differentially methylated sites were found when comparing SMA types or ages at onset in patients with three or four *SMN2* copies separately (**Fig. S9C-F**). In addition, we tested the differentially methylated CpG sites from previous work [19] specifically, but did not replicate any of these associations in our cohort (**Fig. S10**). Finally, we compared changes in the commonly used Hammersmith functional motor scale (HFMSE) scores of a cohort of patients with three or four *SMN2* copies before and 1.5 years after nusinersen treatment initiation (dHFMSE, n=111 **Fig. 4E**). Based on dHFMSE, patients were divided in three treatment response groups: decline, stabilization, and increase. PCA did not show clear clustering of these groups (**Fig. 4F**), and no CpG sites were significantly associated with dHFMSE (**Fig. 4G**). These observations do not completely exclude the possibility that in individual or rare changes in DNA methylation may still affect SMA outcomes. However, the absence of associations between variation in *SMN2* DNA methylation and disease severity or treatment response in these analyses excludes *SMN2* DNA methylation as an epigenetic biomarker of SMA in blood.

**Figure 4:**
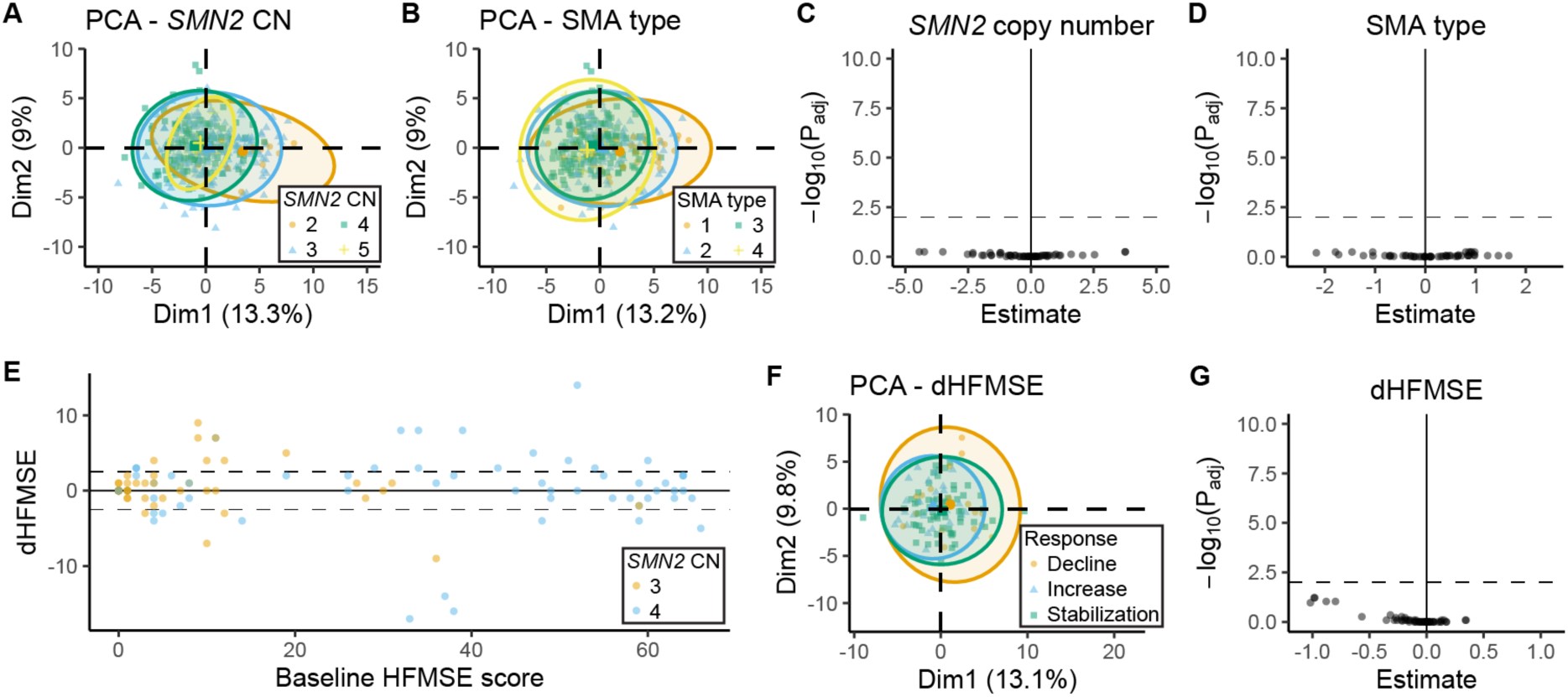
No association was found between *SMN2* DNA methylation and *SMN2* copy number, SMA type or treatment response in SMA patients. (A-B) Principal component analysis (PCA) of DNA methylation on *SMN2* determined by bisulfite sequencing, colored by *SMN2* copy number (A) or SMA type (B). (C-D) Differential methylation analysis for *SMN2* copy number (n=359) (C) and SMA type (n=352) (D). No differentially methylated sites were found (p_adj_<0.01). (E) Difference in HFMSE score (dHFMSE) 1.5 years after nusinersen treatment versus before nusinersen treatment in patients with three or four copies of *SMN2*, plotted against baseline HFMSE score. (E) PCA of DNA methylation on *SMN2* determined by bisulfite sequencing, colored by treatment response group based on HFMSE after nusinersen treatment. Treatment response was divided as follows: Decrease: dHFMSE≤-3; Stabilization: −3<dHFMSE<3; Increase: dHFMSE≤3. (E) Differential methylation analysis for dHFMSE shown with volcano plots (n=111). No differentially methylated sites were found (p_adj_<0.01).

## Discussion

An important outstanding question in the SMA field is the absence of a clear correlation between phenotype and genotype, even among patients with equal *SMN2* copy number. Variation in DNA methylation of *SMN2* has long been hypothesized as a possible explanation for this discrepancy [7,11,13–16]. Indeed, several previous studies suggested that variation in DNA methylation could be a potential modifier of SMA [18,19], but these studies were limited by technological possibilities and sample size, and comprehensive analyses of DNA methylation across the complete *SMN2* gene in large SMA patient cohorts had not been done previously. In this study, we addressed this issue by combining long-read nanopore sequencing to first discover variability in DNA methylation across the complete *SMN2* gene in a limited group of patients, followed by targeted bisulfite sequencing across the identified sites in a large cohort of 365 SMA patients. We identified variable DNA methylation at multiple sites, including the *SMN2* 3’UTR and the intronic transcription site of lncRNA *SMN-AS1*. DNA methylation was significantly associated with age at all tested sites in intron 1 and the 3’ UTR of *SMN2*, and two CpG sites in the promoter and three CpG sites in the transcription site of lncRNA *SMN-AS1*. However, DNA methylation was not significantly associated with *SMN2* copy number, disease severity, treatment response, or *SMN2-FL* mRNA expression. In summary, our results exclude variation in DNA methylation in *SMN2* as an epigenetic biomarker of clinical SMA outcomes but highlight DNA methylation as a possible modifier of SMN expression.

We did not identify methylation differences in the *SMN2* promoter between SMA types in patients with three copies of *SMN2* as reported previously [19]. This previously reported difference may have been observed due to a lower sample size (n=35 versus n=211 in our study) or lenient multiple testing correction due to inclusion of a much smaller number of CpG sites. Another previous study specifically investigated DNA methylation in a group of patients with two *SMN2* copies and either SMA type 1 or SMA type 3. We were unable to replicate these results as the number of patients with two *SMN2* copies in our cohort was limited. In addition, the combination of two *SMN2* copies and SMA type 3 is extremely uncommon, and it cannot be ruled out that the patients included in the previous study were carriers of rare positive modifiers such as c.859G>C or c.835-44A>G, which were not commonly characterized at the time that study was performed. We did not find association between DNA methylation and disease severity in blood as accessible biomarker tissue, but DNA methylation may still play a role in disease-relevant inaccessible tissues such as the spinal cord. Although we did not find DNA methylation in blood measured before treatment initiation to be associated with treatment response in this study, there is still the possibility that epigenetic marks might be involved in the regulation of exon 7 inclusion during treatment [33]. Therefore, longitudinal epigenetic studies may still hold promise for identifying additional targets to strengthen the ability of splice modifiers to increase SMN expression.

In contrast to limited variability in DNA methylation in the *SMN2* promoter region, we observed significant variation in the *SMN2* 3’UTR. Previous studies have associated 3’UTR methylation of other genes with both increased gene expression [34–36], decreased gene expression [37–42], or both [43,44]. Therefore, it is likely that 3’UTR DNA methylation can affect gene expression in different ways for different genes, and through different mechanisms, such as altered protein binding [45], alternative polyadenylation [46,47] or alternative splicing [41]. In the *SMN* genes specifically, Marasco *et al.* showed that a nusinersen-like ASO promotes chromatin-silencing mark H3K9me2, slowing down RNA polymerase II elongation and inhibiting exon 7 inclusion [33]. Reduced DNA methylation in gene bodies is linked to increased chromatin accessibility [48], and increased DNA methylation and H3K9 methylation with heterochromatin [49]. Therefore, reduced DNA methylation at the 3’ end *SMN2* could potentially be associated with faster RNA polymerase II elongation and increased exon 7 inclusion. Although we identified no significant association between *SMN2-FL* expression and DNA methylation in blood, further association analyses between DNA methylation and (long-read) RNA sequencing in other tissues may indicate whether DNA methylation affects splicing and expression of specific exons and isoforms of the *SMN* gene.

DNA methylation is associated with age across mammalian tissues, and this association can be either positive or negative, varying per CpG site [50]. Our current study identified several CpG sites, mostly located in intron 1 and the 3’ UTR of *SMN2,* that were associated with age in our patient cohort. For most sites, we observed that higher age was associated with reduced levels of DNA methylation. A similar pattern was found in the *TARDBP* gene in human motor cortex of healthy subjects and ALS patients, and accelerated DNA demethylation in ALS was associated with earlier disease onset [41]. Therefore, it would be interesting to determine whether age-related decrease of DNA methylation in *SMN2* is also present in healthy subjects. Analyzing this in large control cohorts, however, is challenging as short-read sequencing of bisulfite converted DNA samples does not allow to distinguish *SMN1* and *SMN2* in persons that carry both genes in contrast to SMA patients who only carry copies of *SMN2*. This may become possible in the future when long-read sequencing allowing haplotype-based analysis becomes more routine and scalable. Previous research has shown that SMN protein levels decline rapidly during the perinatal period in *post-mortem* spinal cord [9]. Similarly, *SMN* mRNA and SMN protein expression have been reported to decrease with age in peripheral blood mononuclear cells [10] but remain relatively stable in primary fibroblasts derived from adult patients [10,51]. These findings highlight the potential differences in SMN requirements between young children and adult patients, with potential implications for treatment requirements. While infants and young children may benefit from therapies that maximize SMN expression especially during the first few months of life, this may change in adulthood when patients may have different needs in terms of maintaining sufficient SMN levels as they age. The age-related changes in *SMN2* methylation patterns observed in our study could represent an important consideration for the development of epigenetic therapies that aim to modulate SMN expression across the lifespan. Although speculative, strategies based on the use of catalytically inactive Cas9 (dCas9) fused to the catalytic domain of TET1 (TET1CD) for demethylation [52] or the catalytic domain of DNA methyltransferase 3A (DNMT3A) to increase methylation [53], could form a potential epigenetic therapeutic target for SMA.

## Conclusions

We used nanopore long-read sequencing to reveal extensive variation in DNA methylation across the *SMN2* gene, with low methylation levels observed not only in the promoter region but also in the transcription site of *SMN-AS1*, the 3’ UTR, and various intronic regions. With targeted bisulfite sequencing of 365 SMA patients, no associations were found between DNA methylation and sex, *SMN2* copy number, disease severity or treatment response, excluding DNA methylation in *SMN2* as an epigenetic biomarker of clinical SMA outcomes. We identified 22 CpG sites across *SMN2* where DNA methylation was significantly associated with age, highlighting DNA methylation as a possible modifier of SMN expression during development and aging.

## Methods

### Study population

We included 370 SMA patients from our single-center prevalence cohort study in the Netherlands, as detailed in **Table 1 and Table S1-3**; 24 patients overlap between nanopore and bisulfite sequencing datasets. The study protocol (09307/NL29692.041.09) was approved by the Medical Ethical Committee of the University Medical Center Utrecht and was registered in the Dutch registry for clinical studies and trials (https://www.ccmo.nl/). Written informed consent was obtained from all adult participants, as well as from parents or guardians for patients under 18 years of age. *SMN1*, *SMN2*, and *NAIP* copy numbers were quantified using multiplex ligation-dependent probe amplification (MLPA) (MRC Holland, SALSA MLPA Probemix P021 SMA Version B1) following the manufacturer’s instructions (https://www.mrcholland.com/). The clinical classification of SMA type was conducted based on motor milestones and age at onset as previously described, with type 1 as non-sitters, type 2 as sitters, and type 3 and 4 as walkers [54]. To capture the broad genotypic and phenotypic diversity within the SMA population, the study included patients with *SMN2* copy numbers ranging from two to five, and SMA types ranging from 1b to 4. Whole blood samples were collected in EDTA tubes for DNA extraction, and 3 mm dermal biopsies were obtained for the generation of primary fibroblasts. For 111 patients with three or four *SMN2* copies and receiving nusinersen treatment, Hammersmith Functional Motor Scale – Expanded (HFMSE) scores were available that had been determined before treatment start and 1.5 years after treatment initiation. For PCA, we stratified patients based on treatment response: decrease for dHFMSE≤-3; stabilization for −3<dHFMSE<3; increase for dHFMSE≤3 [55].

### DNA extraction and bisulfite conversion

For bisulfite sequencing, DNA was extracted from whole EDTA blood with the Bl chemagic DNA Blood 4k kit (Revvity, CMG-1074). DNA concentration was quantified with the Quant-iT™ 1X dsDNA BR Assay (Invitrogen, Q33267). Genomic Quality Number (GQN) was determined with the Fragment Analyzer Genomic DNA 50kb Kit (Agilent, DNF-467). 500ng of DNA was bisulfite-converted using the EZ-96 DNA Methylation-Lightning Kit (Zymo, D5033) according to the manufacturer’s instructions. Concentration of converted DNA was measured with a spectrophotometer (Thermo Scientific, Nanodrop 2000) with the ssDNA setting.

### Polymerase chain reaction for bisulfite amplicon sequencing

Primers to amplify the top strand of bisulfite-converted DNA were designed with the Zymo Bisulfite Primer Seeker (https://zymoresearch.eu/pages/bisulfite-primer-seeker) with default options and intended product sizes of 450-550bp. Primers included a sequencing adapter-compatible overhang (5’-TCGTCGGCAGCGTCAGATGTGTATAAGAGACAG-3’ to 5’ end of the forward primer, 5’-GTCTCGTGGGCTCGGAGATGTGTATAAGAGACAG-3’ to 5’ end of the reverse primer). Primer sequences (Integrated DNA Technologies) and amplicon-specific conditions are listed in **Table S4**. Eighteen PCRs were performed on each bisulfite-converted patient DNA sample. Each 10uL PCR reaction contained 2.4μL PCR-grade water, 5μL KAPA HiFi HotStart Uracil+ ReadyMix (2X) (Roche, 7959052001), 0.3μL of each primer (10μM), and a variable amount of DNA optimized for each reaction (**Table S4**). The PCR was run using a Biorad T100 thermocycler (#1861096) with the following protocol: 95°C for 3 minutes; 36 cycles of 98°C for 30 seconds, annealing at variable temperatures for 15 seconds (ramp rate 0.5°C/s), followed by 72°C for 15 seconds (ramp rate 1.1°C/s); and a final elongation at 72°C for 1 minute. All 18 amplicons were visualized on a 2% agarose gel containing Sybr Safe (Fisher Scientific, #10328162) for a subset of eight randomly selected samples and imaged on a Biorad ChemiDoc™ MP Imaging System (#12003154). Band intensity was quantified with Fiji software (version 1.54). Relative band intensity was used for determining the pooling ratio for amplicon pooling, e.g. a relative brightness of 2x resulted in a pooling ratio of 0.5x. All 18 amplicons were pooled per patient and subsequently cleaned with AMPure XP beads at a ratio of 0.65x to eliminate PCR reagents and small DNA fragments. A subset of samples was run on the Agilent 2200 TapeStation (#G2965A) with Agilent High Sensitivity D1000 ScreenTape (5067-5584) and High Sensitivity D1000 Reagents (5067-5585) to check for overamplification.

### Library preparation and Illumina sequencing

A second PCR was performed on each patient sample containing 18 pooled amplicons to attach Illumina DNA/RNA UD Indexes Sets A-D (Illumina, 20091654, 20091656, 20091658, 20091660) barcodes to the compatible overhangs that were included in the first PCR (see above). Each 25uL PCR reaction contained 5.5μL PCR-grade water, 12.5μL NEBNext Q5 Hot Start HiFi PCR Master Mix (NEB, M0544L), 2.5μL of unique Illumina® DNA/RNA UD Index, and 2.5ng of DNA in a volume of 2μL. The PCR was run in a Biorad T100 thermocycler with the following steps: 98°C for 30 seconds; six cycles of 98°C for 10 seconds followed by 65°C for 75 seconds; and a final elongation at 65°C for five minutes.

The PCR products were purified with AMPure XP beads at a ratio of 0.8x. A subset of samples was measured on the Agilent 2200 TapeStation with Agilent High Sensitivity D1000 ScreenTape and High Sensitivity D1000 Reagents to check if the Illumina DNA/RNA UD Indexes had successfully annealed. The concentration of a subset of individual samples was measured with the Qubit dsDNA High Sensitivity kit (Thermo Fisher Scientific, #Q32854) kit to check if concentrations were roughly equal, and all samples were pooled at equal volume. The DNA concentration of the final pooled library was measured with the Qubit dsDNA High Sensitivity kit. 785pM library with 2% PhiX Sequencing Control was loaded onto an Illumina Nextseq2000 P1 flow cell and sequenced with 2×300bp paired-end read settings.

### Nanopore data processing

Nanopore sequencing data was generated previously (**Table S1**)[23]; one patient was excluded due to a partial deletion of *SMN1* exon 1-6, another patient was excluded due to unavailability of clinical data. In summary, raw sequencing data was basecalled with Guppy v6.1.2 with the SUP model (dna_r9.4.1_450bps_modbases_5mc_cg_sup.cfg) and mapped to the T2T-CHM13 reference genome masked for a ∼170kb region surrounding *SMN2* [23]. If data from both blood and fibroblasts of the same patient was available, this data was not merged. Polyploid haplotype phasing was performed with GATK and WhatsHap as described previously [23]. Methylation calling was performed for full bam files and per-haplotype bam files with modbam2bed v1.0 with options -e -m 5mC -r chr5:71375000-71425000 --cpg. Methylation calls from forward and reverse strands were merged per CpG site. Methylation percentage was calculated from modbam2bed output with the formula: Nmod / (Nmod + Ncan) * 100.

### Illumina data processing

Bisulfite sequencing data was processed with the methylseq v2.6.0 workflow from nf-core [56], including raw data QC with FastQC, adapter sequence and quality trimming (Phred<20) with Trim Galore!, read alignment to the masked T2T-CHM13 reference genome [23] with Bismark and extraction of methylation calls with Bismark. Default options were used, except for: --unmapped, --skip_deduplication and --save_align_intermeds. For each sample, the bismark.cov.gz file from the methylation_coverage output directory was loaded into R4.4.0 [57] and the data for all samples were merged into one dataset. Methylation percentage was used directly from the cov.gz file. The data was filtered to only contain CpG sites that are located on the intended amplicons and had a read depth (methylated reads + unmethylated reads) of 100x or more. When the C or G of a CpG site was a known SNV position [25], it was removed, to prevent any SNVs to be mistakenly interpreted as methylation changes, since unconverted DNA was not sequenced. Additional data filtering was applied before statistical testing with linear models, to reduce dimensionality: sites with little variation between samples (methylation percentage standard deviation <5%) were removed, and highly correlated sites with Spearman’s R>0.9 were treated as one site by averaging (**Fig. S3**).

### Visualization and exploration of methylation data

Methylation percentages were visualized per sample or per haplotype using the pheatmap v1.0.12 package [58] in R v4.4.0. Row clustering was performed with the ward.D2 method and Euclidian distance. PCA was performed with the function PCA from the FactoMineR v2.11 package [59] followed by the function fviz_pca_ind from package factoextra v1.0.7 [60].

### DNA motif analyses

The sequence surrounding the tested CpG sites (20bp upstream and 20bp downstream) was extracted with bedtools v2.30.0. DNA binding motifs for CTCF were downloaded from CTCF_HUMAN.H11MO.0.A on hocomoco11.autosome.org [31]. FIMO (https://meme-suite.org/meme/doc/fimo.html [32]) was used to scan the *SMN1/2* gene sequence for CTCF binding motifs. Sequence logos were generated using https://weblogo.threeplusone.com/create.cgi [61].

### Statistics

Required sample size was calculated in R4.4.0 using the pwr v1.3-0 package [62] using the pwr.f2.test function. To detect a medium effect size (Cohen’s f^2^=0.15) [63] in a linear model with a power of 80%, alpha of 0.01 and assuming 4 regression degrees of freedom (corresponding to three covariates), a sample size of 115 is required. Differential methylation analysis was performed using linear models in R4.4.0 with the following formula [64]: dependent_variable ∼ independent_variable + covariates; using methylation percentage as dependent variable. For each independent variable (such as age at sampling or SMA type), one formula was made per CpG site, thus one p-value was calculated for every CpG site and corrected for multiple testing with the false discovery rate (FDR) method. Covariates used were age at sampling in years (except when age was independent variable) [50,65], sex [66], library size per *SMN* copy and GQN. Continuous independent variables such as age at sampling were included in the model as numeric values, whereas factors with two levels were converted to 0 and 1. Ordinal independent variables such as *SMN2* and *NAIP* copy number were used as numeric values. SMA type was converted to numeric in the following way before using it as independent variable: type 1 to 0, type 2 to 1, type 3 to 2 and type 4 to 3. For testing concordance in ONT data, the data was divided into two groups due to small sample size: less severe with three copies of *SMN2* and type 2b or less severe and with four copies of *SMN2* and type 3b or less severe; more severe with three copies of *SMN2* and type 2a or more severe and with four copies of *SMN2* and type 3a or more severe. Similarly, we compared the less severe group versus the concordant and more severe group taken together. Volcano plots were made by plotting the -log10 of the adjusted p-value against the estimate of the regression coefficient belonging to the independent variable for each CpG site. Results of all statistical tests including effect sizes for all CpG sites are denoted in **Table S5**.

## Declarations

### Ethics approval and consent to participate

The study protocol (09307/NL29692.041.09) was approved by the Medical Ethical Committee of the University Medical Center Utrecht and registered at the Dutch registry for clinical studies and trials (https://www.ccmo.nl/). Written informed consent was obtained from all adult patients, and from patients and/or parents additionally in case of children younger than 18 years old.

## Consent for publication

Not applicable.

### Declaration of interests

WvR has sponsored research agreements with Biogen and Astra Zeneca. JHV reports to have sponsored research agreements with Biogen, Eli Lilly and Astra Zeneca. The other authors declare no competing interests.

## Data Availability

The datasets used and/or analyzed during the current study are available from the corresponding author on reasonable request but are not publicly available due to privacy restrictions. The code used for analyses in this study is available at: https://github.com/ghaaften/SMA_DNA_methylation.

## Acknowledgements

We thank the patients who participated in this study. This work was supported by grants from Stichting Spieren voor Spieren (to WLvdP), the European Union’s Horizon 2020 Research and Innovation Program under the Marie Skłodowska-Curie grant (H2020 Marie Skłodowska-Curie Actions) agreement no. 956185 (SMABEYOND ITN to WLvdP, EJNG) and Prinses Beatrix Spierfonds (W.OB21-01 to EJNG). This project has received funding from the European Research Council (ERC) under the European Union’s Horizon 2020 research and innovation programme (grant agreement n° 772376 - EScORIAL). We acknowledge the Utrecht Sequencing Facility (USEQ) for providing sequencing service and data. USEQ is subsidized by the University Medical Center Utrecht and The Netherlands X-omics Initiative (NWO project 184.034.019).

## Author contributions

Conceptualization, MMZ, MGE, DG, EJNG, GWvH; experimental studies, MMZ, IS, JK, JG, RG; bioinformatics and data analysis MMZ, MGE, DG, HWMvD, JvdS, RAJZ, PJH, WvR; clinical data & patient material, FA, RIW, WLP; writing–original draft, MMZ, EJNG, GWvH; writing–review, all; resources, EJNG, WLvdP; supervision, EJNG, JHV, WLvdP, GWvH; funding acquisition, EJNG, GWvH, WLvdP.

## Notes

### Author Declarations

The Medical Ethical Committee of the University Medical Center Utrecht gave ethical approval for this work

