## Supplemental information for "Comprehensive analysis across *SMN2* excludes DNA methylation as an epigenetic biomarker for spinal muscular atrophy"

Contents

- Supplemental tables 1-3
- Supplemental figures 1-10

Supplemental files

- Table_S4_bisulfite_amplicon_overview.xlsx (Overview of bisulfite sequencing amplicons, including PCR conditions)
- Table_S5_differential_methylation_test_results.xslx (Results of all statistical tests performed for differential methylation analyses)

**Supplemental table 1:** Baseline characteristics of SMA patients sequenced with Nanopore sequencing.

| | Blood | | Fibroblasts | | | |
|---|---|---|---|---|---|---|
| | **3x*SMN2*** | **4x*SMN2*** | **2x*SMN2*** | **3x*SMN2*** | **4x*SMN2*** | **5x*SMN2*** |
| **Total** | 6 | 4 | 3 | 10 | 8 | 1 |
| **Sex** | | | | | | |
| Male | 3 (50.0%) | 2 (50.0%) | 1 (33.3%) | 6 (60.0%) | 7 (87.5%) | 1 (100%) |
| Female | 3 (50.0%) | 2 (50.0%) | 2* (66.7%) | 4 (40.0%) | 1 (12.5%) | 0 (0%) |
| **SMA type** | | | | | | |
| Type 1b | 0 (0%) | 0 (0%) | 2 (66.7%) | 0 (0%) | 0 (0%) | 0 (0%) |
| Type 1c | 2 (33.3%) | 0 (0%) | 0 (0%) | 1 (10.0%) | 0 (0%) | 0 (0%) |
| Type 2a | 2 (33.3%) | 0 (0%) | 0 (0%) | 6 (60.0%) | 1 (12.5%) | 0 (0%) |
| Type 2b | 1 (16.7%) | 2 (50.0%) | 0 (0%) | 1 (10.0%) | 1 (12.5%) | 0 (0%) |
| Type 3a | 1 (16.7%) | 0 (0%) | 1* (33.3%) | 1 (10.0%) | 1 (12.5%) | 0 (0%) |
| Type 3b | 0 (0%) | 1 (25.0%) | 0 (0%) | 1 (10.0%) | 3 (37.5%) | 1 (100%) |
| Type 4 | 0 (0%) | 1 (25.0%) | 0 (0%) | 0 (0%) | 2 (25.0%) | 0 (0%) |
| **Age at onset (years)** | | | | | | |
| Mean (SD) | 0.708 (0.244) | 7.00 (9.11) | 0.403 (0.528) | 1.77 (2.75) | 11.0 (9.03) | 15.5 (NA) |
| Median [Min, Max] | 0.646 [0.417, 1.00] | 3.00 [1.50, 20.5] | 0.208 [0, 1.00] | 0.813 [0.500, 9.50] | 12.7 [0.833, 24.5] | 15.5 [15.5, 15.5] |
| **Age at sampling (years)** | | | | | | |
| Mean (SD) | 12.3 (14.4) | 45.1 (13.6) | 11.4 (17.7) | 11.7 (18.3) | 35.0 (24.3) | 21.1 (NA) |
| Median [Min, Max] | 6.71 [4.50, 41.5] | 45.3 [30.0, 59.7] | 1.92 [0.417, 31.8] | 6.58 [2.08, 63.3] | 31.6 [9.92, 70.1] | 21.1 [21.1, 21.1] |

*Includes one patient with one *SMN1* gene copy with a pathogenic mutation.

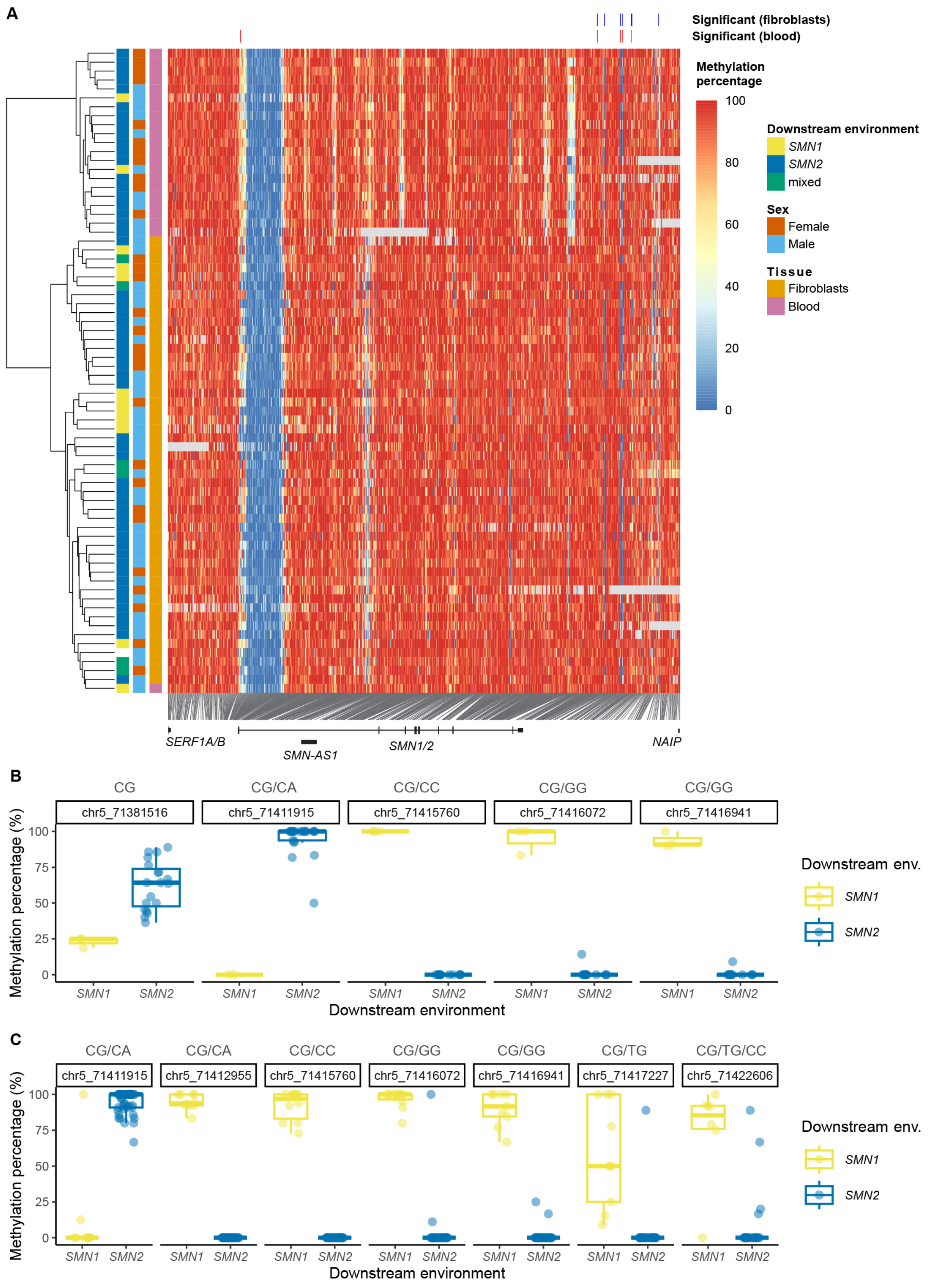


**Supplemental figure 1:** Per-haplotype analysis of DNA methylation in nanopore sequencing data of SMA patients.

(A) Heatmap of DNA methylation per haplotype, determined by nanopore sequencing at T2T-CHM13 coordinates chr5:71375000-71425000, containing 710 CpG sites. 72 out of 108 haplotypes with less than 25% NA are included. Hierarchical clustering was performed according to the ‘ward.D2’ method. Sites that were differentially methylated between haplotypes with downstream *SMN1* versus *SMN2* environment (Fig. 1H-I) are shown at the top of the figure.

(B-C) Methylation percentage at CpG sites that were differentially methylated between different downstream environments in blood (B) and fibroblasts (C). Many of these sites are known SNV sites (Zwartkruis et al. 2024); the possible nucleotide changes are indicated above each site.

**Supplemental table 2:** Baseline characteristics of SMA patients sequenced with bisulfite sequencing, with *SMN1* with a known pathogenic mutation.

| | **2x*SMN2* (N=2)** | **3x*SMN2* (N=2)** | **Overall (N=4)** |
|---|---|---|---|
| **Sex** | | | |
| Male | 1 (50.0%) | 1 (50.0%) | 2 (50.0%) |
| Female | 1 (50.0%) | 1 (50.0%) | 2 (50.0%) |
| **SMA type** | | | |
| Type 1 | 0 (0%) | 1 (50.0%) | 1 (25.0%) |
| Type 3 | 2 (100%) | 1 (50.0%) | 3 (75.0%) |
| **Age at onset (years)** | | | |
| Mean (SD) | 2.50 (2.12) | 1.00 (0.707) | 1.75 (1.55) |
| Median [Min, Max] | 2.50 [1.00, 4.00] | 1.00 [0.500, 1.50] | 1.25 [0.500, 4.00] |
| **Age at sampling (years)** | | | |
| Mean (SD) | 40.4 (14.1) | 30.8 (42.9) | 35.6 (26.7) |
| Median [Min, Max] | 40.4 [30.4, 50.4] | 30.8 [0.474, 61.2] | 40.4 [0.474, 61.2] |

**Supplemental table 3:** Baseline characteristics of SMA patients sequenced with bisulfite sequencing, with a known c.859G>C variant in *SMN2*.

| | **2x*SMN2* (N=2)** | **Overall (N=2)** |
|---|---|---|
| **Sex** | | |
| Male | 1 (50.0%) | 1 (50.0%) |
| Female | 1 (50.0%) | 1 (50.0%) |
| **SMA type** | | |
| Type 2 | 1 (50.0%) | 1 (50.0%) |
| Type 3 | 1 (50.0%) | 1 (50.0%) |
| **Age at onset (years)** | | |
| Mean (SD) | 4.83 (5.89) | 4.83 (5.89) |
| Median [Min, Max] | 4.83 [0.667, 9.00] | 4.83 [0.667, 9.00] |
| **Age at sampling (years)** | | |
| Mean (SD) | 36.7 (17.9) | 36.7 (17.9) |
| Median [Min, Max] | 36.7 [24.1, 49.3] | 36.7 [24.1, 49.3] |

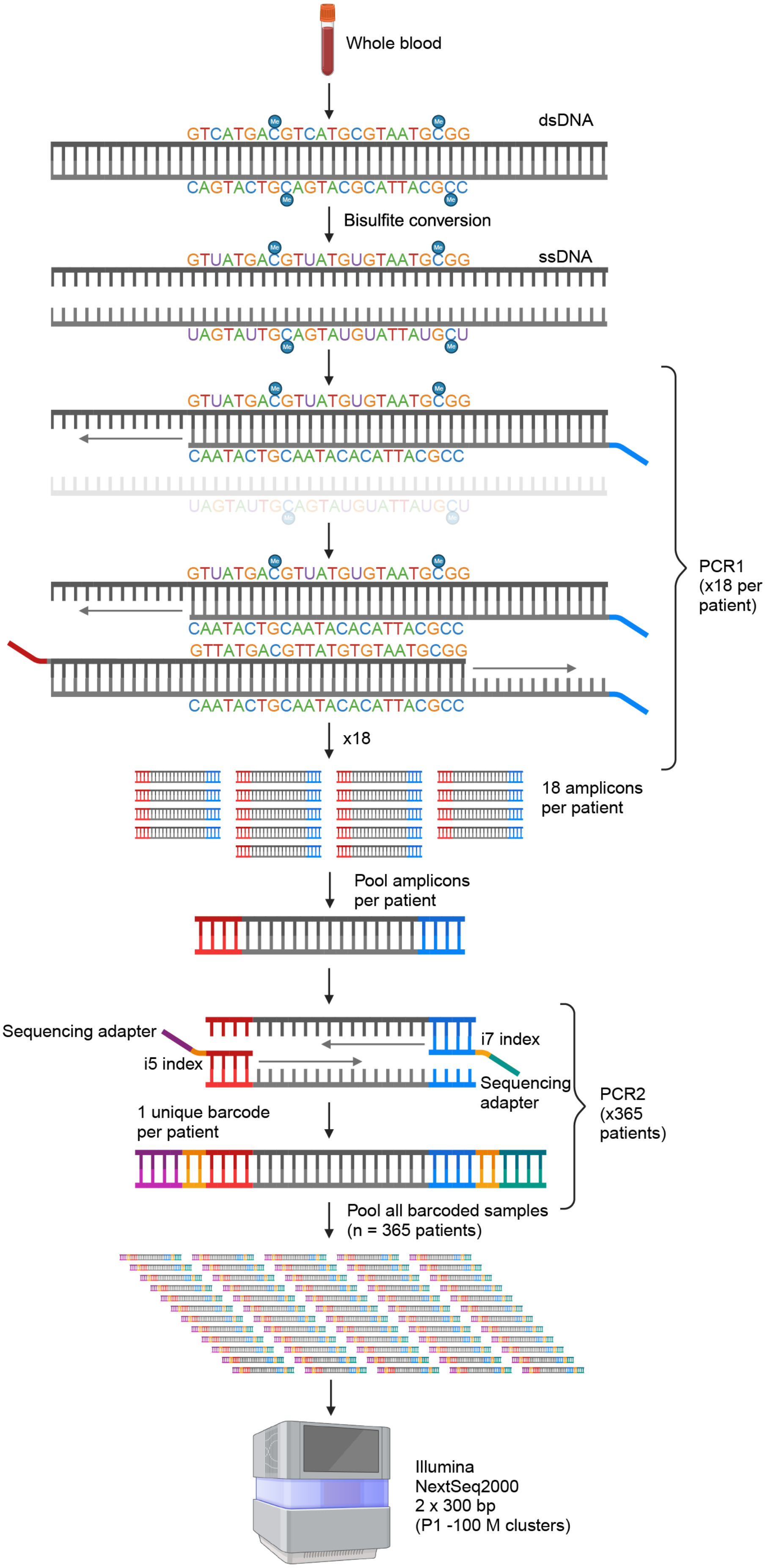
Whole blood
dsDNA
GTCATGACGTCATGCGTAATGCGG
CAGTACTGCAGTACGCATTACGCC
Bisulfite conversion
ssDNA
GTUATGACGTUATGUGTAATGCGG
UAGTAUTGCAGTAUGUATTAUGCU
CAATACTGCAATACACATTACGCC
GTTATGACGTTATGTGTAATGCGG
PCR1
(x18 per
patient)
x18
18 amplicons
per patient
Pool amplicons
per patient
Sequencing adapter
i5 index
i7 index
Sequencing
adapter
PCR2
(x365
patients)
1 unique barcode
per patient
Pool all barcoded samples
(n = 365 patients)
Illumina
NextSeq2000
2 x 300 bp
(P1 -100 M clusters)

**Supplemental figure 2**: Bisulfite sequencing methods. Double-stranded DNA was isolated from whole blood and bisulfite-converted, resulting in single-stranded DNA. The top strand of the converted DNA was amplified by PCR, using primers with an overhang compatible with the sequencing adapters (Illumina DNA/RNA UD Indexes). 18 such PCRs were performed per patient and all amplicons were pooled. For each patient, sequencing adapters were ligated to the amplicons in a second round of PCR. Samples from 365 patients were pooled and sequenced on an Illumina NextSeq2000. Created with Biorender.com.

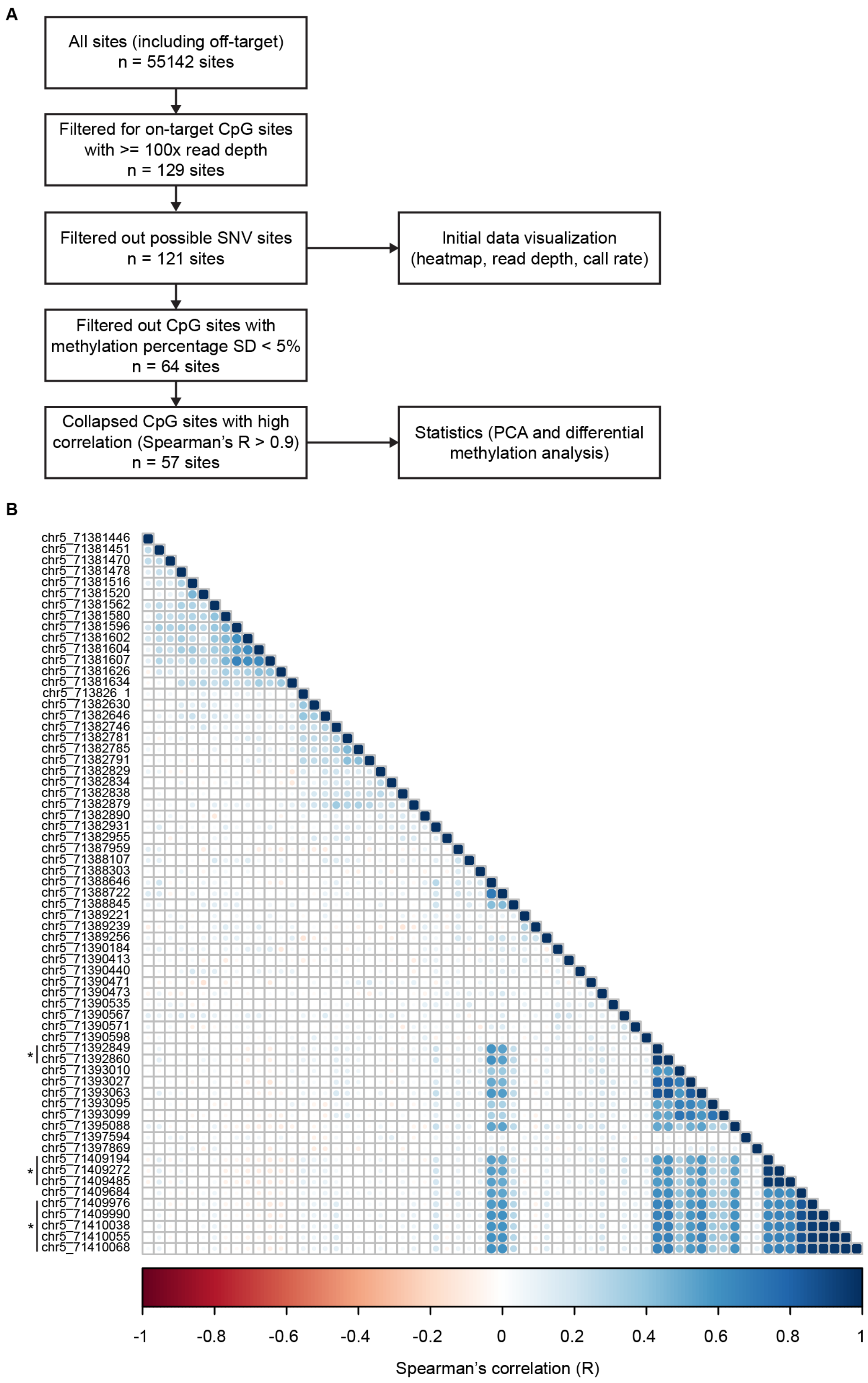


**Supplemental figure 3:** Dimensionality reduction by filtering of CpG sites.

(A) CpG sites were filtered for mapping on the intended targets, a minimum read depth of 100x, not being a known SNV site (Chen et al. 2023) and not having a low standard deviation (SD). Lastly, sites with high Spearman correlation (R>0.9) as shown in (B), were condensed into one site by taking their mean.

(B) Correlation plot showing the Spearman correlation between methylation at the tested CpG sites, used for the last filtering step of (A). Sites with Spearman R>0.9 are indicated by an asterisk.

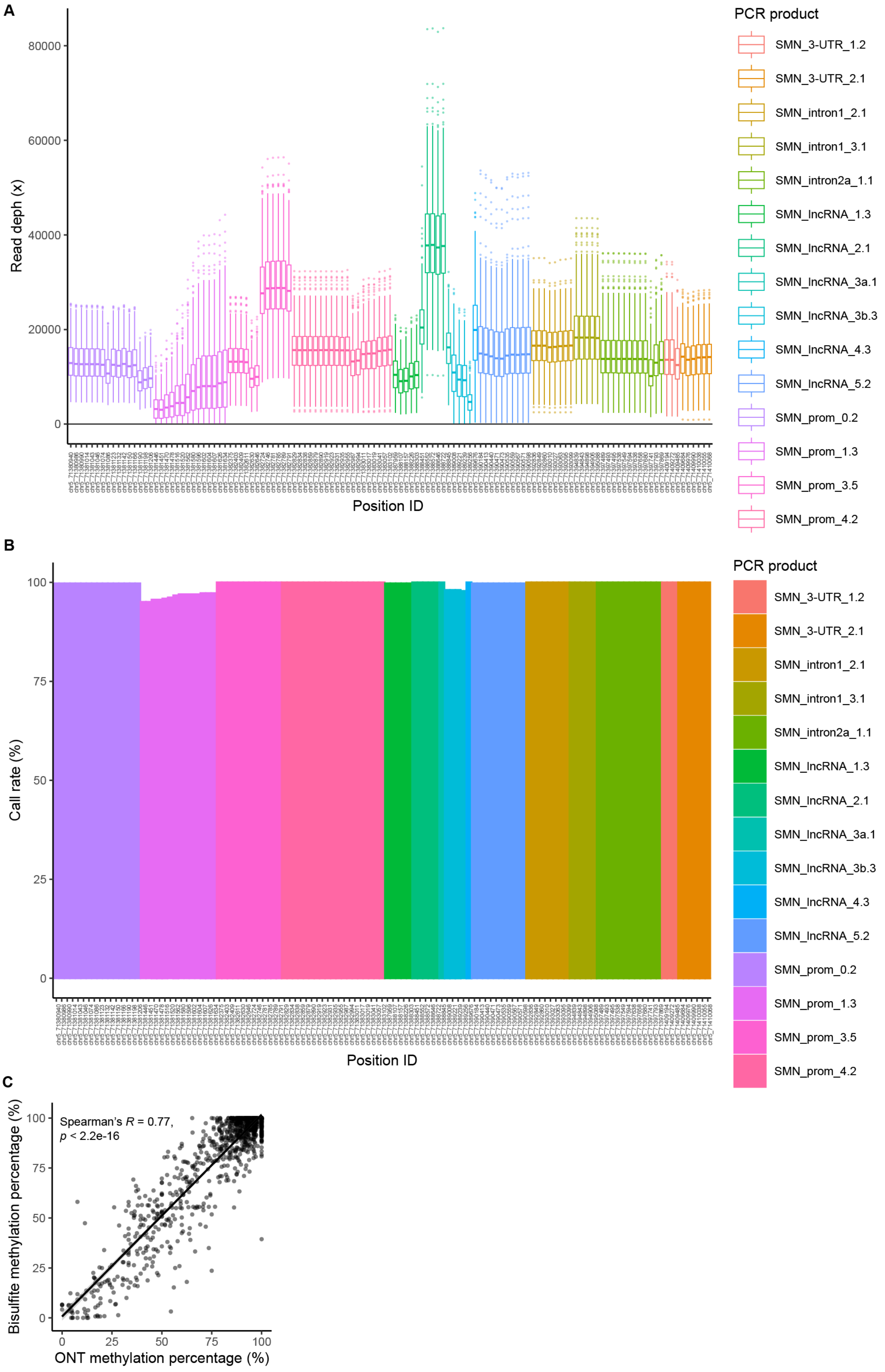
A
Read depth (x)
80000
60000
40000
20000
0
Position ID
PCR product
SMN_3-UTR_1.2
SMN_3-UTR_2.1
SMN_intron1_2.1
SMN_intron1_3.1
SMN_intron2a_1.1
SMN_lncRNA_1.3
SMN_lncRNA_2.1
SMN_lncRNA_3a.1
SMN_lncRNA_3b.3
SMN_lncRNA_4.3
SMN_lncRNA_5.2
SMN_prom_0.2
SMN_prom_1.3
SMN_prom_3.5
SMN_prom_4.2
B
Call rate (%)
100
75
50
25
0
Position ID
PCR product
SMN_3-UTR_1.2
SMN_3-UTR_2.1
SMN_intron1_2.1
SMN_intron1_3.1
SMN_intron2a_1.1
SMN_lncRNA_1.3
SMN_lncRNA_2.1
SMN_lncRNA_3a.1
SMN_lncRNA_3b.3
SMN_lncRNA_4.3
SMN_lncRNA_5.2
SMN_prom_0.2
SMN_prom_1.3
SMN_prom_3.5
SMN_prom_4.2
C
Spearman's R = 0.77,
p < 2.2e-16
Bisulfite methylation percentage (%)
100
75
50
25
0
0
25
50
75
100
ONT methylation percentage (%)

**Supplemental figure 4:** Performance of bisulfite sequencing.

(A) Median read depth per CpG site represented by the midline of boxplots, after filtering for sites with at least 100x read depth and filtering out possible SNV sites. The box represents the interquartile range.

(B) Call rate per CpG site: percentage of samples that have ≥100x read depth, giving a valid methylation call.

(C) Spearman correlation between nanopore and bisulfite methylation percentages, determined from nine samples for which both data types were available; $p<2.2e-16$, Spearman’s $R=0.77$.

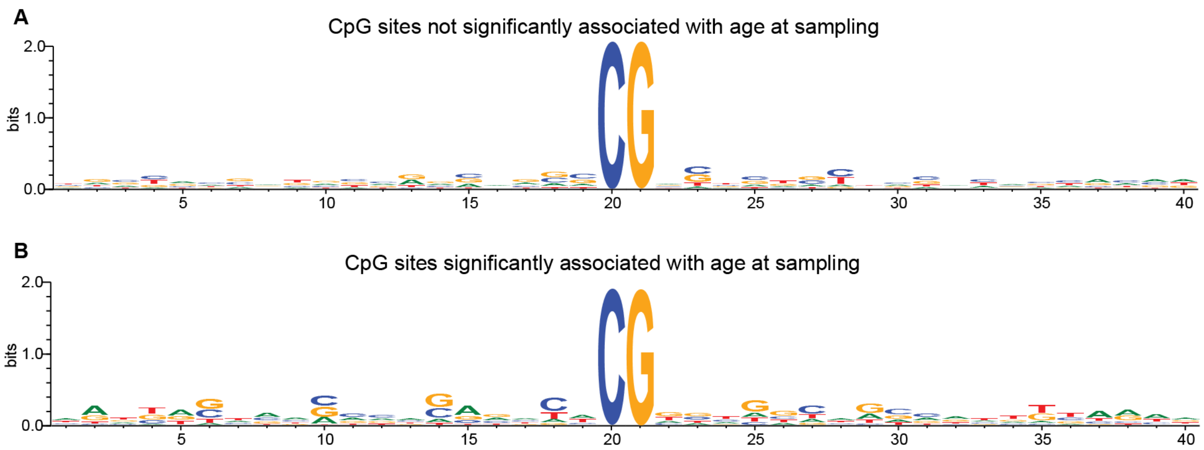


**Supplemental figure 5:** Nucleotide motif surrounding CpG sites that are not significantly associated with age (n=42, A) and CpG sites that are significantly associated with age (n=22, B).

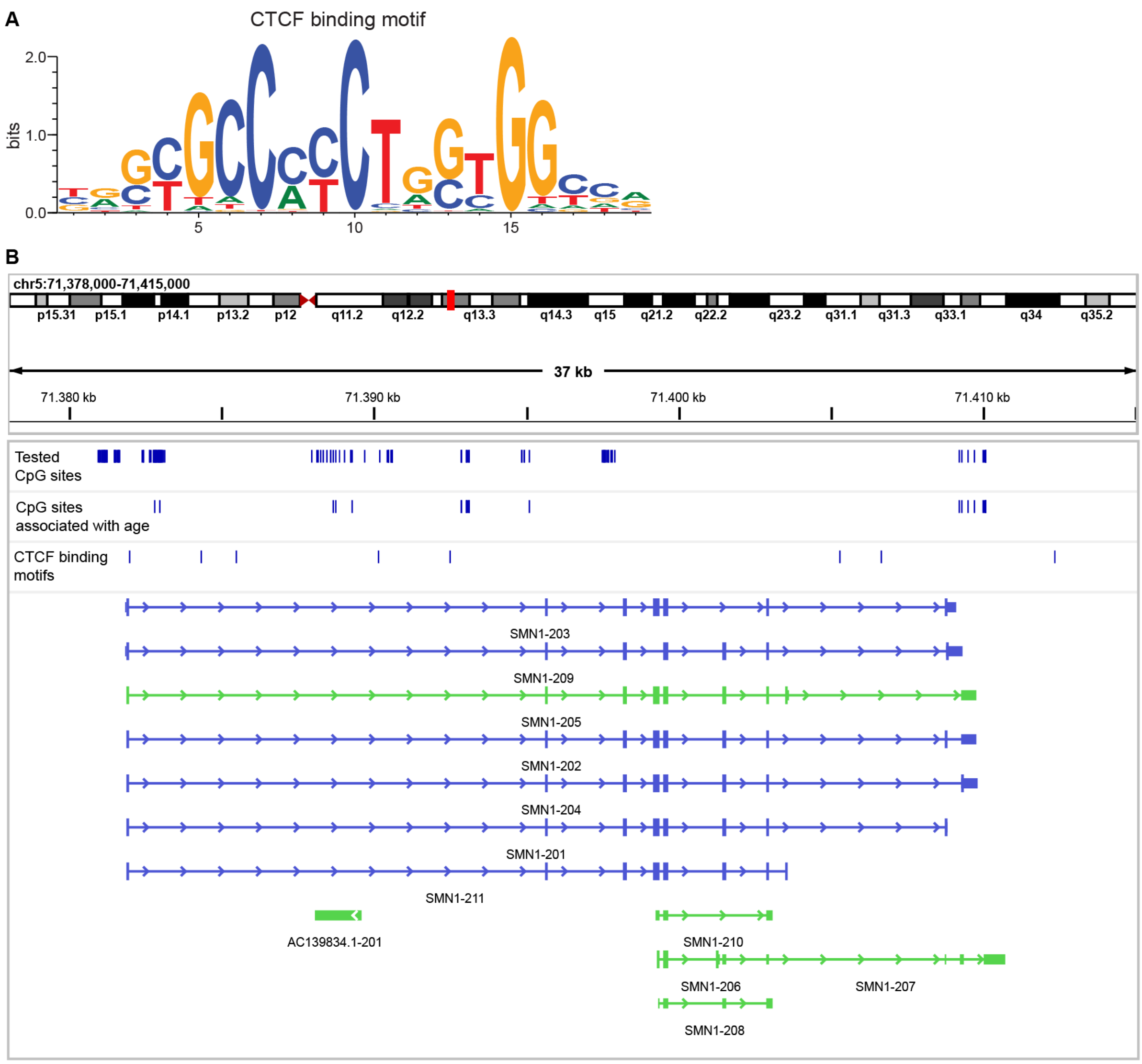


**Supplemental figure 6:** CTCF binding motifs on the *SMN* gene.

(A) CTCF binding motif.

(B) IGV snapshot of the *SMN1* gene on T2T-CHM13 reference genome, *SMN1-202* being the canonical transcript. CTCF binding motifs do not overlap with the CpG sites associated with age.

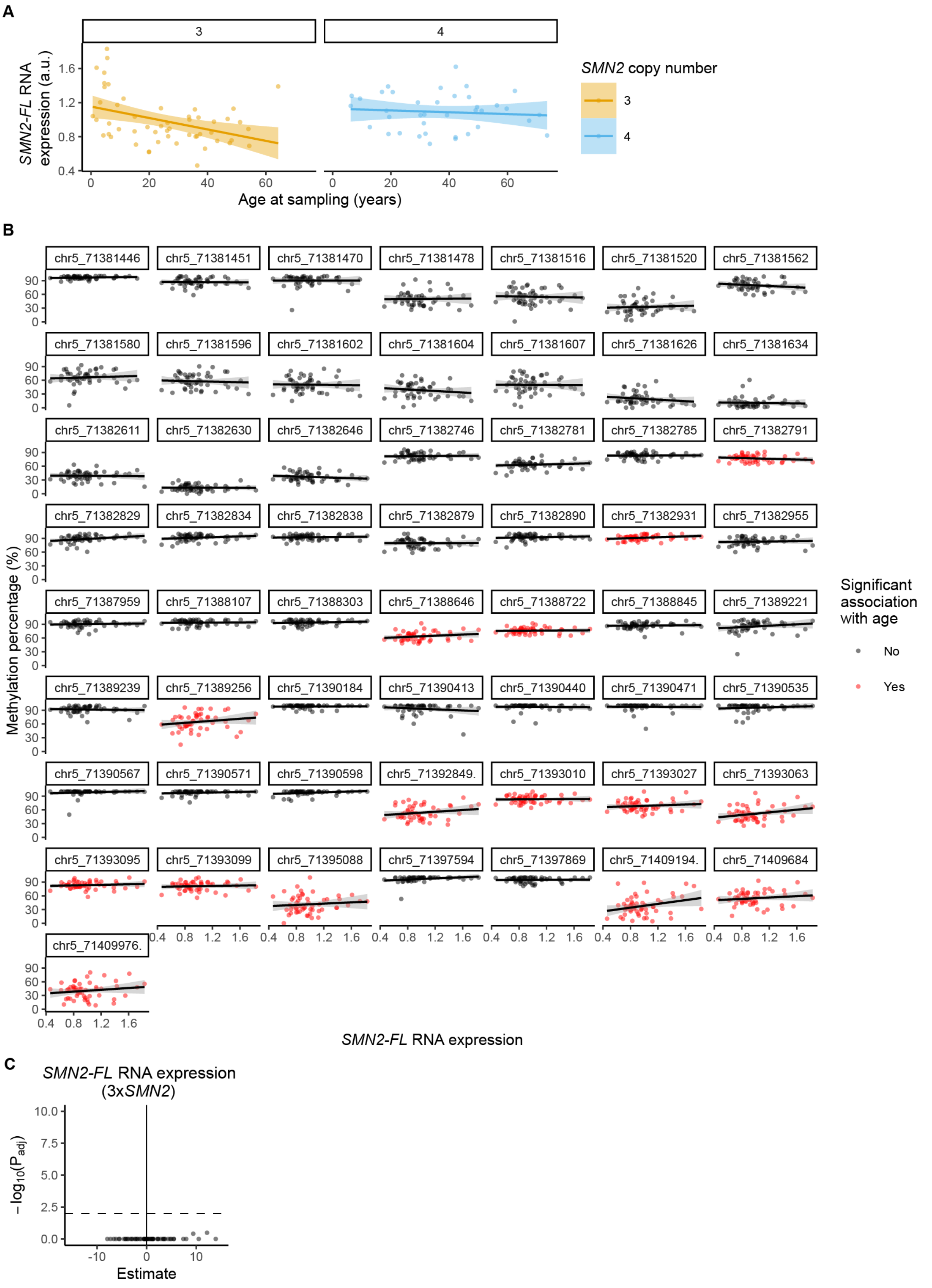


**Supplemental figure 7:** Association between age, *SMN2-FL* RNA expression and DNA methylation in *SMN2*.

(A) *SMN2-FL* RNA expression (Wadman et al. 2020) was associated with age at sampling in patients with three copies of *SMN2* (simple linear regression, used model: SMN2-FL=1.153-0.006714*(age), adjusted R-squared: 0.1407, F(1,51)=9.514, p=0.003289) but not in patients with four copies of *SMN2* (simple linear regression, used model: SMN2-FL=1.129-0.001071*(age), adjusted R-squared: -0.01986, F(1,36)=0.2795, p=0.6003).

(B) DNA methylation percentage plotted against *SMN2-FL* RNA expression for patients with three *SMN2* copies. CpG sites at which DNA methylation is associated with age are shown in red. Collapsed sites are indicated with an asterisk (*).

(C) Differential methylation analysis for different amounts of *SMN2-FL* RNA expression in SMA patients with three *SMN2* copies (n=53) as shown in (B). No CpG sites were significantly associated with *SMN2-FL* RNA expression ($p_{adj}$<0.01).

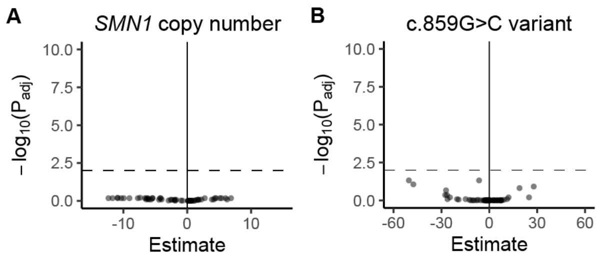


**Supplemental figure 8:** Differential methylation analysis for patients with known functional genetic variants.

(A) Differential methylation analysis for patients with *SMN1* with a pathogenic mutation (n=4) versus patients with a homozygous *SMN1* deletion with similar *SMN2* copy number of two or three (n=230). No CpG sites were significantly associated with presence of the *SMN1* gene ($p_{adj}<0.01$).

(B) Differential methylation analysis for patients with the c.859G>C variant in *SMN2* (n=2) versus patients without this variant and the same *SMN2* copy number of two (n=15). No CpG sites were significantly associated with presence of the c.859G>C variant ($p_{adj}<0.01$).

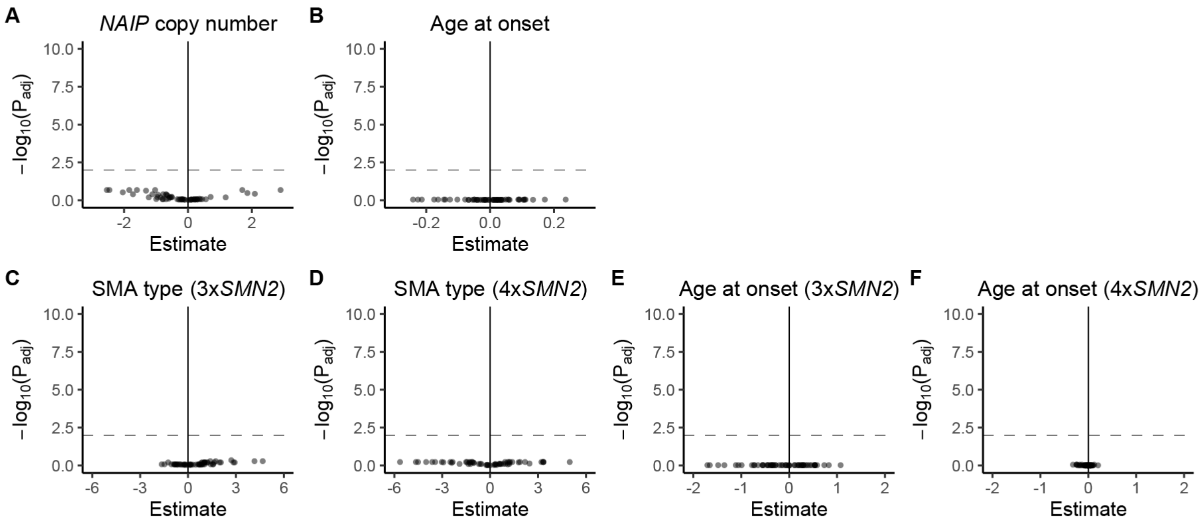


**Supplemental figure 9:** Differential methylation analysis.

(A) Differential methylation analysis between patients with different *NAIP* copy numbers, ranging from zero to four (n=359). No differentially methylated sites were found ($p_{adj}$<0.01).

(B) Differential methylation analysis between patients with different ages at onset, ranging from 0 to 43 years old (n=333). No differentially methylated sites were found ($p_{adj}$<0.01).

(C-D) Differential methylation analysis between different SMA types in patients with three *SMN2* copies (C, n=215) and four *SMN2* copies (D, n=122). No differentially methylated sites were found ($p_{adj}$<0.01).

(E-F) Differential methylation analysis between different ages at onset in patients with three *SMN2* copies (E, n=204) and four *SMN2* copies (F, n=109). No differentially methylated sites were found ($p_{adj}$<0.01).

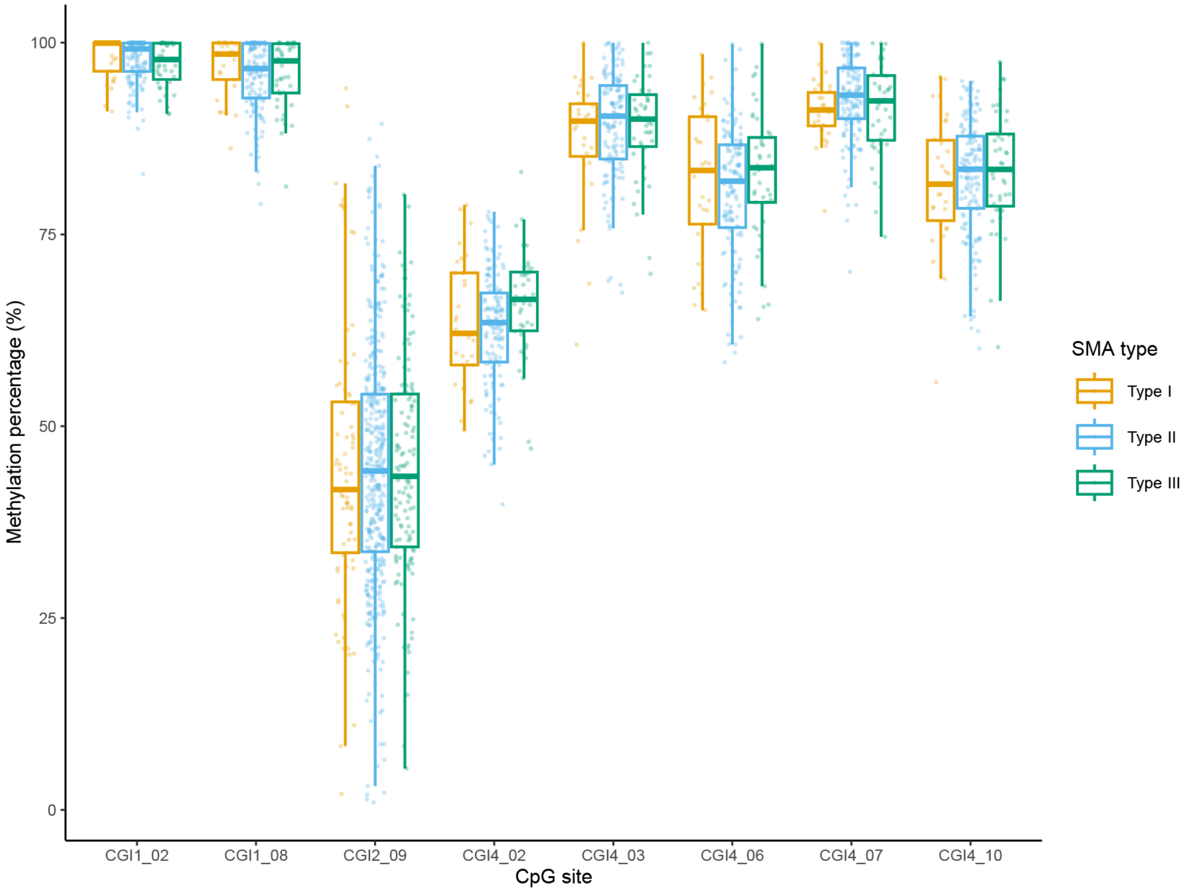


**Supplemental figure 10:** DNA methylation at sites reported to be associated with SMA disease severity in (Cao et al. 2016), in patients with three copies of *SMN2* and SMA type 1 (n=30), type 2 (n=138) and type 3 (n=43). For each CpG site, the Kruskal-Wallis rank sum test was performed with FDR multiple testing correction: CGI1_02 (chr5:71381014): $p_{adj}$=1; CGI1_08 (chr5:71381150): $p_{adj}$=1; CGI2_09 (chr5:71381602, chr5:71381604 and chr5:71381607): $p_{adj}$=1; CGI4_02 (chr5:71382781): $p_{adj}$=0.2804; CGI4_03 (chr5:71382829): $p_{adj}$=1; CGI4_06 (chr5:71382879): $p_{adj}$=1; CGI4_07 (chr5:71382890): $p_{adj}$=0.60656; CGI4_10 (chr5:71382955): $p_{adj}$=1.
